# The impact of London’s Ultra Low Emission Zone on respiratory prescribing: a synthetic control study

**DOI:** 10.64898/2026.08.27.26361515

**Authors:** George Williams, Thomas Allen

## Abstract

Urban air pollution remains a significant public health concern, contributing to premature deaths and adverse health outcomes. However, there is little causal research evaluating the effectiveness of policies designed to improve air quality. This study assesses the impact of all three stages of London’s Ultra Low Emission Zone (ULEZ) on air pollution, via PM_2.5_ levels, and respiratory health, via prescription records for bronchodilator and respiratory corticosteroid medications. Analyses are at general practice level, using a generalised synthetic control method to estimate causal impacts. Stage 1 was associated with a statistically significant but negligible 0.77% reduction in PM_2.5_ levels, with no corresponding change in prescribing. Stage 2 produced a paradoxical 2.69% increase in PM_2.5_, alongside a 4.44% decrease in inhaled corticosteroid quantity but a 12.51% increase in average daily quantity (ADQ) usage, suggesting a worsening of disease severity among existing patients. Stage 3 yielded a 2.69% PM_2.5_ reduction and a modest 2.18% decrease in bronchodilator ADQ usage. Spillover effects beyond the ULEZ boundary were statistically significant, but negligible. We find overall that the ULEZ had minimal effects on both air quality and respiratory prescribing across all three stages. These findings provide new insights into the effectiveness of ULEZ policies in reducing air pollution and its associated health impacts, suggesting the zone’s effects are considerably smaller than previously reported, and that integration with broader policy measures may be necessary to achieve meaningful public health gains.

## 1 Introduction

Vehicle emissions are a major source of pollution, contributing substantially to poor air quality worldwide, especially in urban areas. The health effects of air pollution are well-established; exposure has been found to increase risks of stroke, heart disease, lung cancer, and respiratory disorders [1, 2]. It is estimated that air pollution causes 8.34 million premature deaths annually, disproportionately affecting children, the elderly, and the immunocompromised [3]. In response to persistently high pollution levels, ultra low emission zones (ULEZs) have been introduced in urban areas around the world with the aim of reducing vehicle pollution. This study utilises a method seldom used in this field - the generalised synthetic control - to determine the causal impact of London’s ULEZ on prescribing of respiratory medicines in primary care, offering new evidence on whether such policies translate into measurable health benefits.

The Mayor of London introduced the ULEZ in April 2019, initially in Central London before expanding twice to cover all of Greater London. Building on the earlier Low Emission Zone (LEZ) introduced in 2008, which applied only to commercial vehicles, the ULEZ imposes a daily charge on the most polluting vehicles to discourage their use in densely populated areas. The policy has sparked significant political and economic debate, with ongoing disagreements over its effectiveness and necessity. As one of the world’s largest cities, London’s ULEZ has become a model for similar policies across the UK and globally, making the findings of this study relevant to other regions adopting comparable schemes.

While much of the existing literature on air pollution policy focuses on changes in pollution levels or acute hospital admissions, less is known about the effects on management of chronic conditions in primary care. This study addresses this gap by examining whether the introduction and expansion of the London ULEZ affected GP-level prescribing rates of bronchodilators and inhaled corticosteroids. By applying a generalised synthetic control method, we estimate the policy’s impact across its three major implementation phases (2019, 2021, and 2023), offering new evidence on how clean air policies may affect the burden of respiratory disease in the community. These findings contribute to a more comprehensive understanding of the public health effects of urban air quality interventions.

One key mechanism through which ULEZ policies are hypothesised to improve public health is by reducing exposure to fine particulate matter (PM_2.5_), a pollutant emitted by vehicles and strongly associated with respiratory disease exacerbation, leading to increased medication usage [4]. The London Assembly’s one-year report on the London-wide ULEZ expansion estimates a 31% decrease in vehicle-derived PM_2.5_ levels, as reported from roadside monitoring site data [5]. In this paper, we first corroborate these results using satellite-derived PM_2.5_ estimates mapped to GP practice catchment areas, to estimate the reduction in the 6 months following each stage’s introduction^1^.

If the ULEZ was effective in reducing PM_2.5_ exposure, we would expect to see a reduction in primary care respiratory medicines usage, reflecting the reduced burden of respiratory disease. Using the same generalised synthetic control method, we then estimate the change in bronchodilator and inhaled corticosteroid prescriptions over the same time period following each ULEZ stage.

## 2 Methods

The initial ULEZ was introduced in April 2019 and encompassed only the area of inner London already in the congestion charge zone. The first expansion in October 2021 increased the boundary to the North and South Circular Roads, while the most recent expansion has brought all of Greater London and some areas of surrounding counties within the zone. Figure 1 shows a map of the three stages of the ULEZ, compiled using ArcGIS.

**Figure 1:**
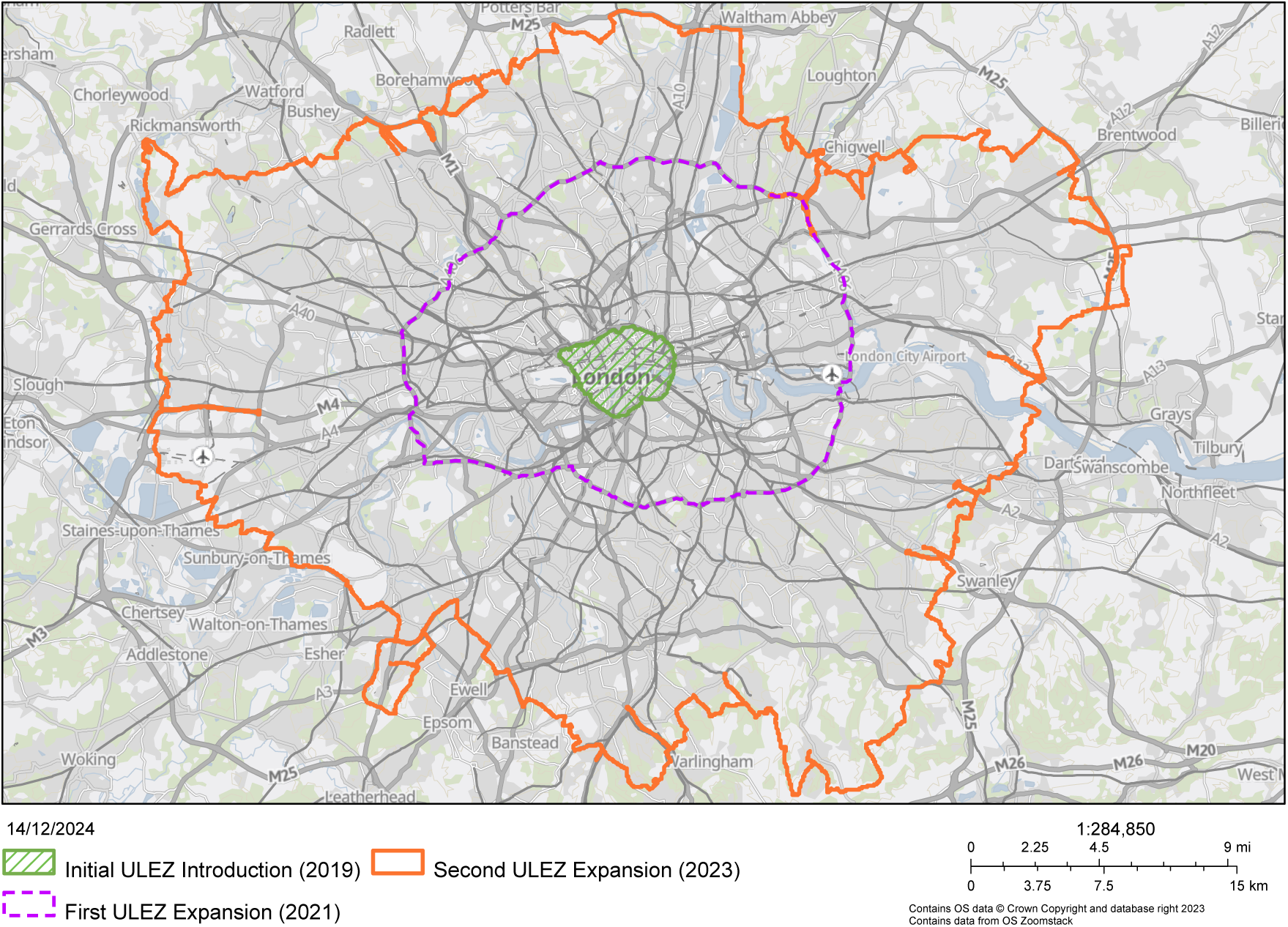
ULEZ Expansion Map by Stage

### 2.1 Study Design

The unit of observation in this study is the GP practice and its surrounding catchment area. As of 2016, there were 7,763 GP practices in England, each serving a defined catchment area within which registered patients live [6]. Data on GP practices, including their addresses, Integrated Care Board (ICB) locations, and population, were sourced from the House of Commons Library and cross-referenced with NHS England datasets. To ensure data consistency over the period, practices that opened after 2016 or were closed at any point during the period of interest were excluded from the analysis; practices that were open but did not report data over the time period were also excluded.

The dataset consists of monthly observations of outcome and control variables from January 2016 to February 2024 inclusive. ULEZ exposure was assigned by matching boundary postcodes to GP practice postcodes. The number of treated practices at each stage is presented in Table 1.

**Table 1:**
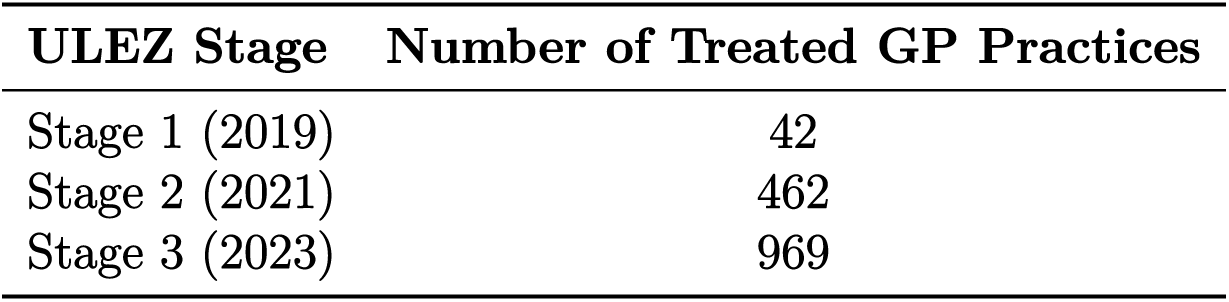
Number of Treated GP Practices in Each ULEZ Stage.

While data was collected for all GP practices, the control group was restricted to a set of 18 cities and large towns^2^, consistent with existing studies [7]. This selection was made to better approximate the characteristics of treated practices while ensuring a diverse demographic profile. The full list of included areas is provided in Table A1.

### 2.2 Outcome Variables

Monthly PM_2.5_ concentration data was obtained at 0.01^◦^*×*0.01^◦^ resolution for all of England from the University of Washington’s regional estimates, provided by the Atmospheric Composition Analysis Group. This dataset provides annual surface PM_2.5_ estimates based on satellite-derived aerosol optical depth, chemical transport models, and ground-based monitoring data, ensuring a comprehensive measurement across the treated and control regions [22]. Using NHS catchment boundary shapefiles, PM_2.5_ values were geographically mapped to each GP practice catchment area using ArcGIS to ensure accurate exposure assignment. While population density estimates were considered, they were found to be insufficiently comprehensive across all treated and control areas. Therefore, we assume uniform population density within each catchment area.

To assess respiratory health, we measure specifically bronchodilators and inhaled corticosteroids, sections 3.1 and 3.2 in the British National Formulary (BNF). Full lists of the medicines in these sections are provided in Tables A2 and A3. Practice-level prescribing data is available at monthly intervals from the NHS English Prescribing Dataset (EPD). Total quantity and average daily quantity (ADQ) usage data for BNF sections 3.1 and 3.2 were obtained from the EPD for each practice over the period. ADQ usage and its use in this study is explained in 2.3.

### 2.3 ADQ Usage Calculation

When estimating changes in respiratory health through prescription analysis, examining quantity alone is insufficient to capture shifts in prescribing activity. For instance, if ULEZ improves respiratory health, as hypothesised, patients may experience symptom relief that justifies switching to a less potent drug. This represents an improvement in health but is not reflected in raw prescription quantity data, as the number of prescriptions issued has remained the same.

To address this, ADQ usage is used as an analytical aid. ADQ is an analytical unit representing the average daily dose of a drug, as predefined by the British National Formulary (BNF) - values for which are provided in Tables A2 and A3.

From this, ADQ usage is calculated as:

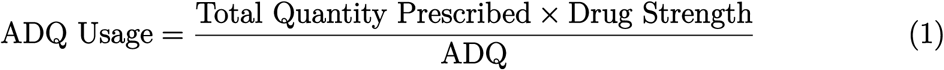

By estimating ADQ usage, we account for changes in the strength of prescribed drugs, not just their volume, providing a more comprehensive view of prescribing trends.

### 2.4 Control Variables

Data on each practice’s demographic profile was sourced from Public Health England’s ‘Fingertips’ dataset. This includes information on ethnicity, sex, and five-year age bands, as well as Index of Multiple Deprivation (IMD) scores. Additionally, practice-level survey data provides estimates of the percentage of active and former smokers—an important covariate in this study, given the well-established link between smoking and cardiorespiratory disease [8, 9]. Data estimating the prevalence of asthma and COPD sufferers in each GP practice were also sourced from the NHS Quality and Outcomes Framework (QOF) dataset [10]. Monthly surface temperature, surface pressure, windspeed, humidity and precipitation data were obtained at 1 km *×* 1 km resolution for all of England from the Met Office’s Had-UK Grid dataset. As pollutant levels are known to be affected by weather patterns [11], these data were included as control variables for regressions. As with PM_2.5_, the grid values were again mapped to each GP practice catchment area to provide monthly weather data for each unit.

### 2.5 Statistical Methods

Whilst analyses have been carried out estimating the effect of low emission zones on air pollution, few studies have employed advanced techniques to accurately elicit the causal effect of the policy on pollution levels or medication usage. The most similar study found upon reviewing the literature was Beshir and Fichera [7]’s difference-in difference (DiD) analysis, using roadside monitoring station data compared with a control comprised of other UK cities. Under the assumption of parallel trends, this approach performs well, however an initial event study of our data suggested London follows a greater downward trend in both pollution and respiratory prescribing levels as compared to our basket of control cities. As such, a DiD would not be appropriate for our study, and would obfuscate the causal impact of the policy.

The empirical strategy proposed in this study is a type of synthetic control method (SCM), allowing an estimation to be made in the presence of non-parallel trends by constructing a weighted combination of control units that best resembles the treated units prior to intervention. Developed from the original method proposed by Abadie and Gardeazabal [12], we use the generalised synthetic control method (GSCM) developed by Xu [13]. This allows for the analysis of panel data with multiple treated units, imputing a counterfactual for each GP practice, as opposed to just London in its entirety. A primer on the GSCM and the rationale for employing it in this study is provided in Appendix A.2.

## 3 Results

Data for all practices was sourced in the first instance, and was filtered to a set of 2,200 practices for analysis, as detailed in the flow chart below.

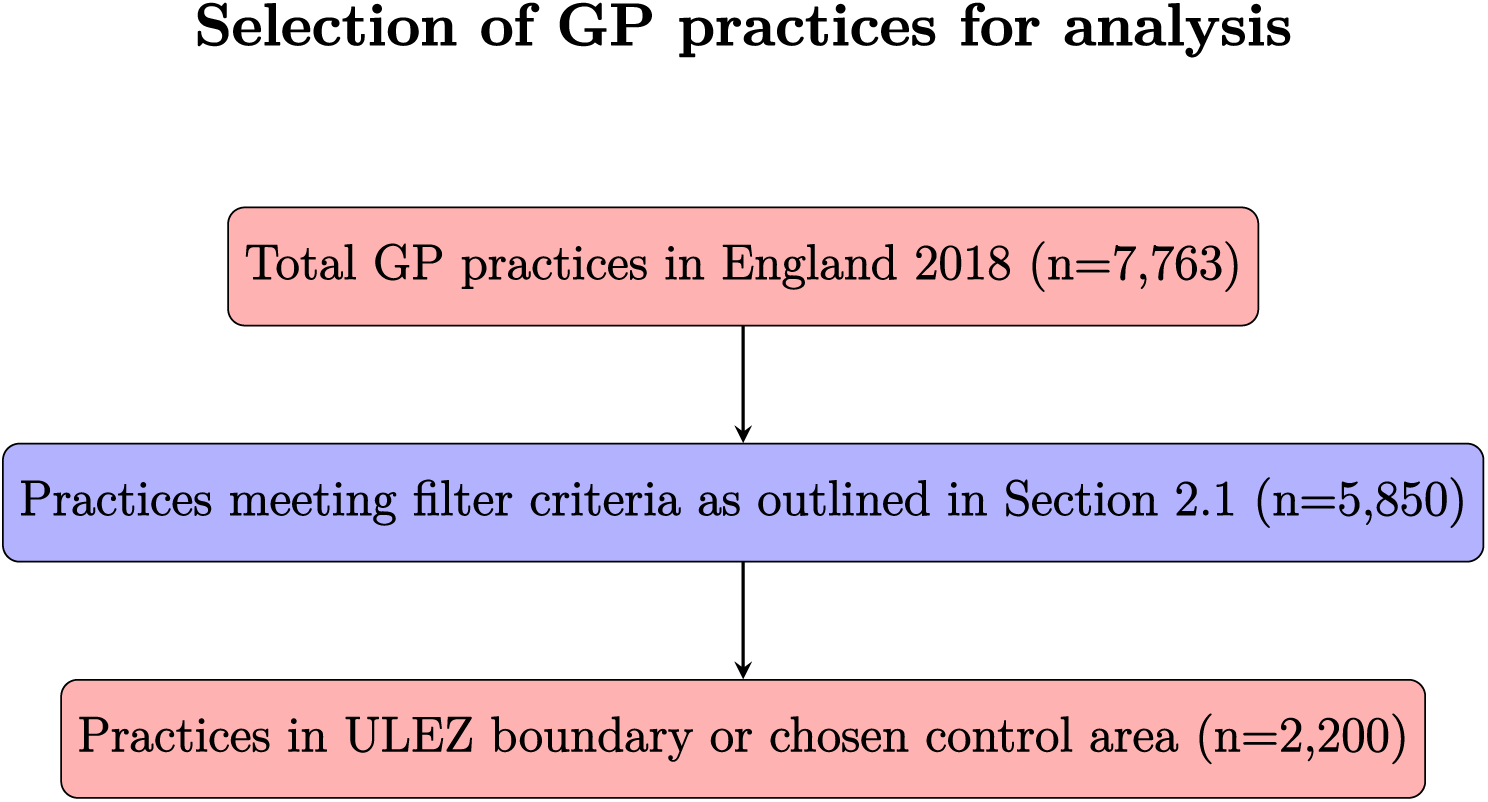

### 3.1 PM_2.5_ Levels

Applying the GCSM method outlined in Section 2.5, we estimate PM_2.5_ counterfactuals. Figures 2, 3, and 4 show the estimated PM_2.5_ change for the treated versus the constructed synthetic control for the first 6 months after each ULEZ stage.

**Figure 2:**
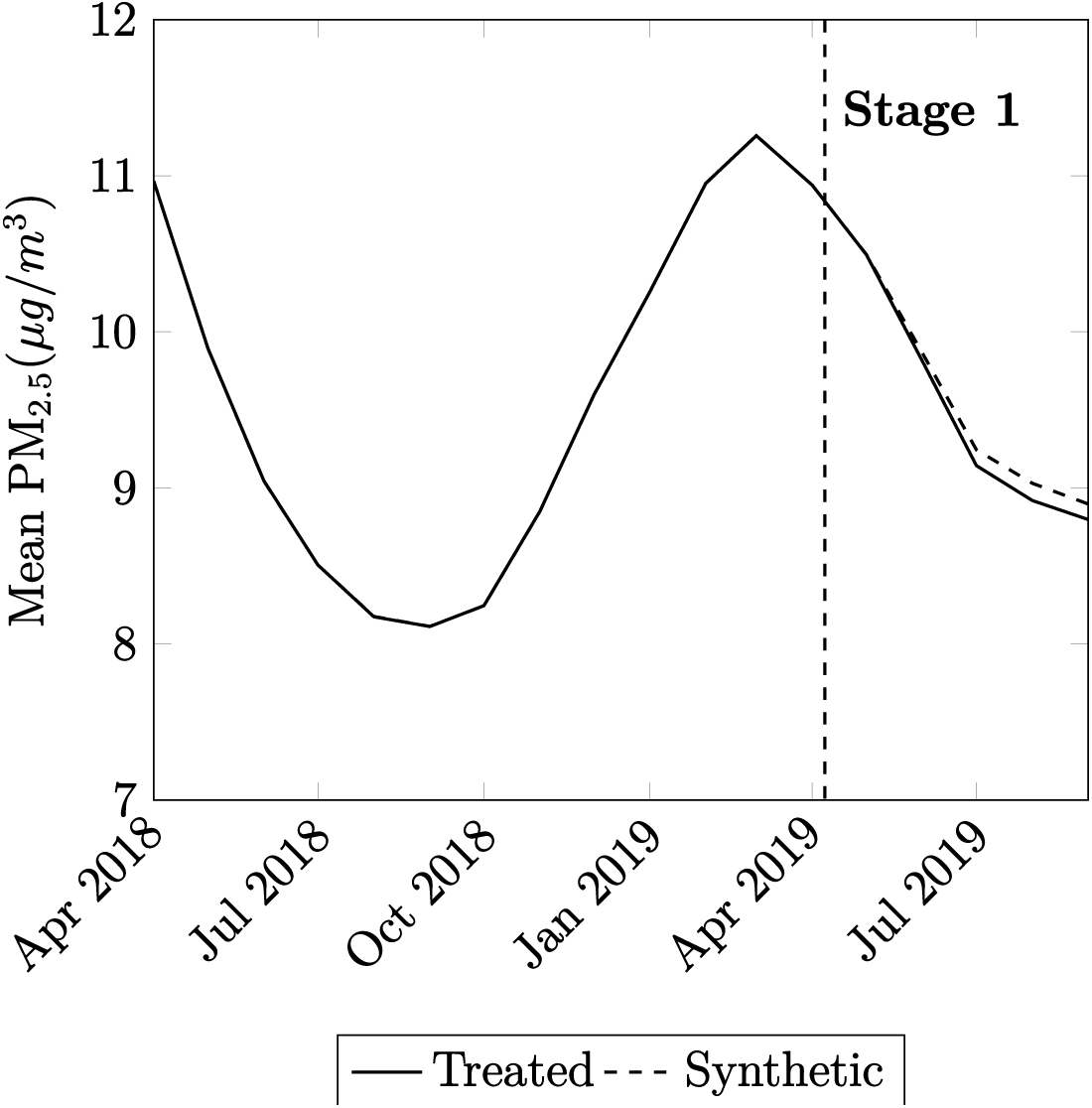
Average PM_2.5_ levels from April 2018 to September 2019 in Treated Group and Synthetic Counterfactual.

**Figure 3:**
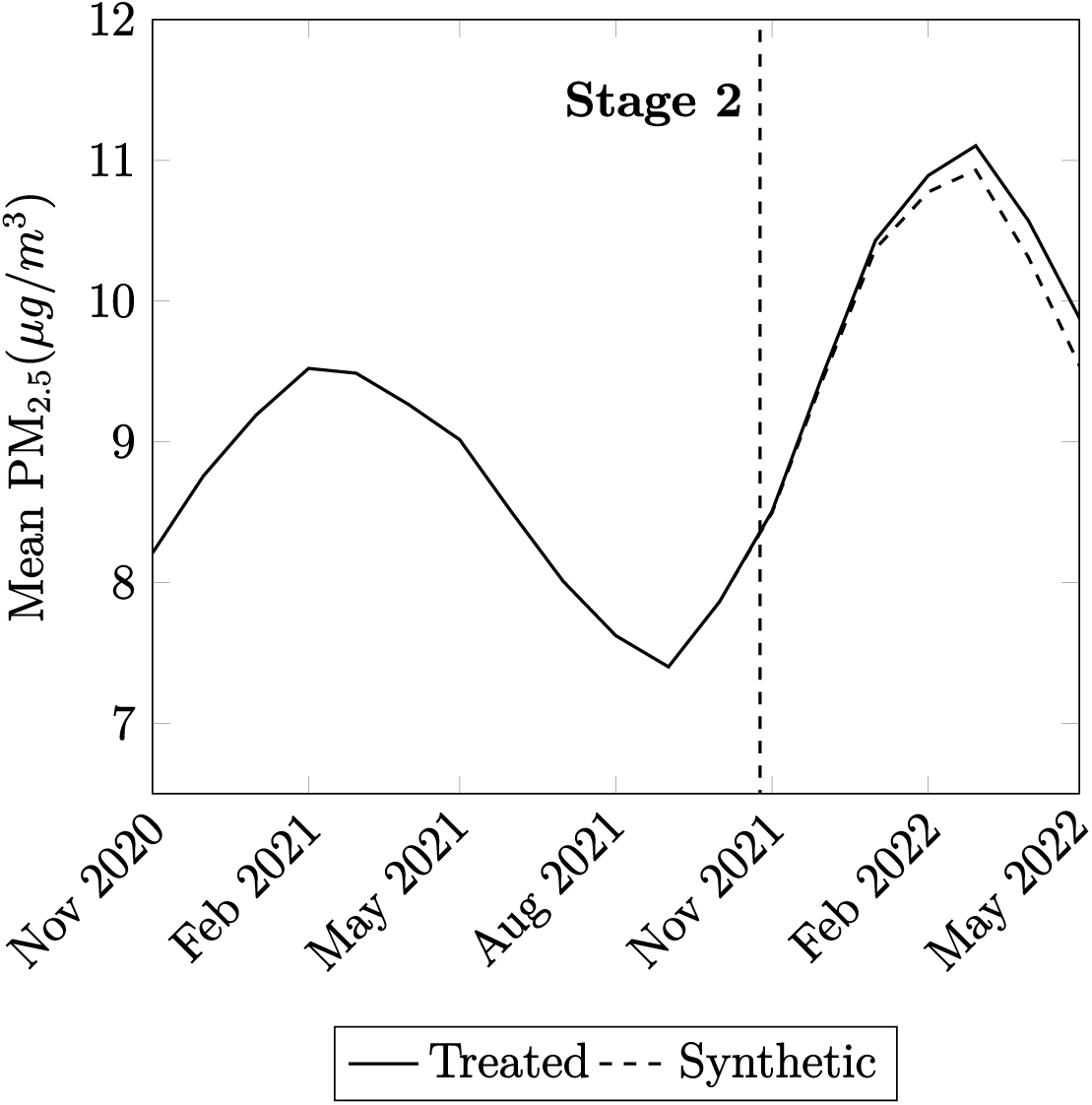
Average PM_2.5_ levels from November 2020 to May 2022 in Treated Group and Synthetic Counterfactual.

**Figure 4:**
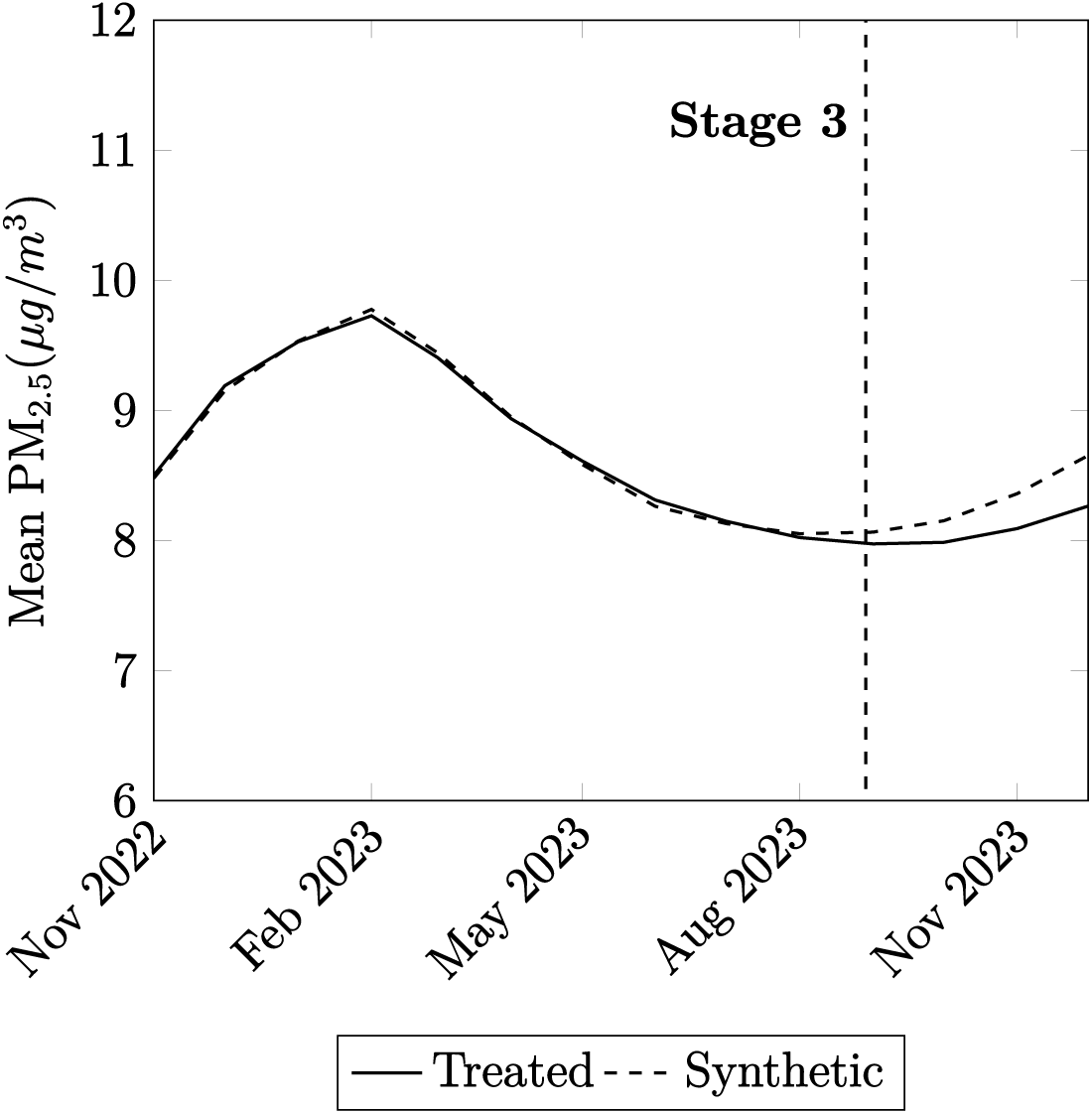
Average PM_2.5_ levels from November 2022 to December 2023 in Treated Group and Synthetic Counterfactual.

Table 2 displays the results shown in the figures above.

**Table 2:**
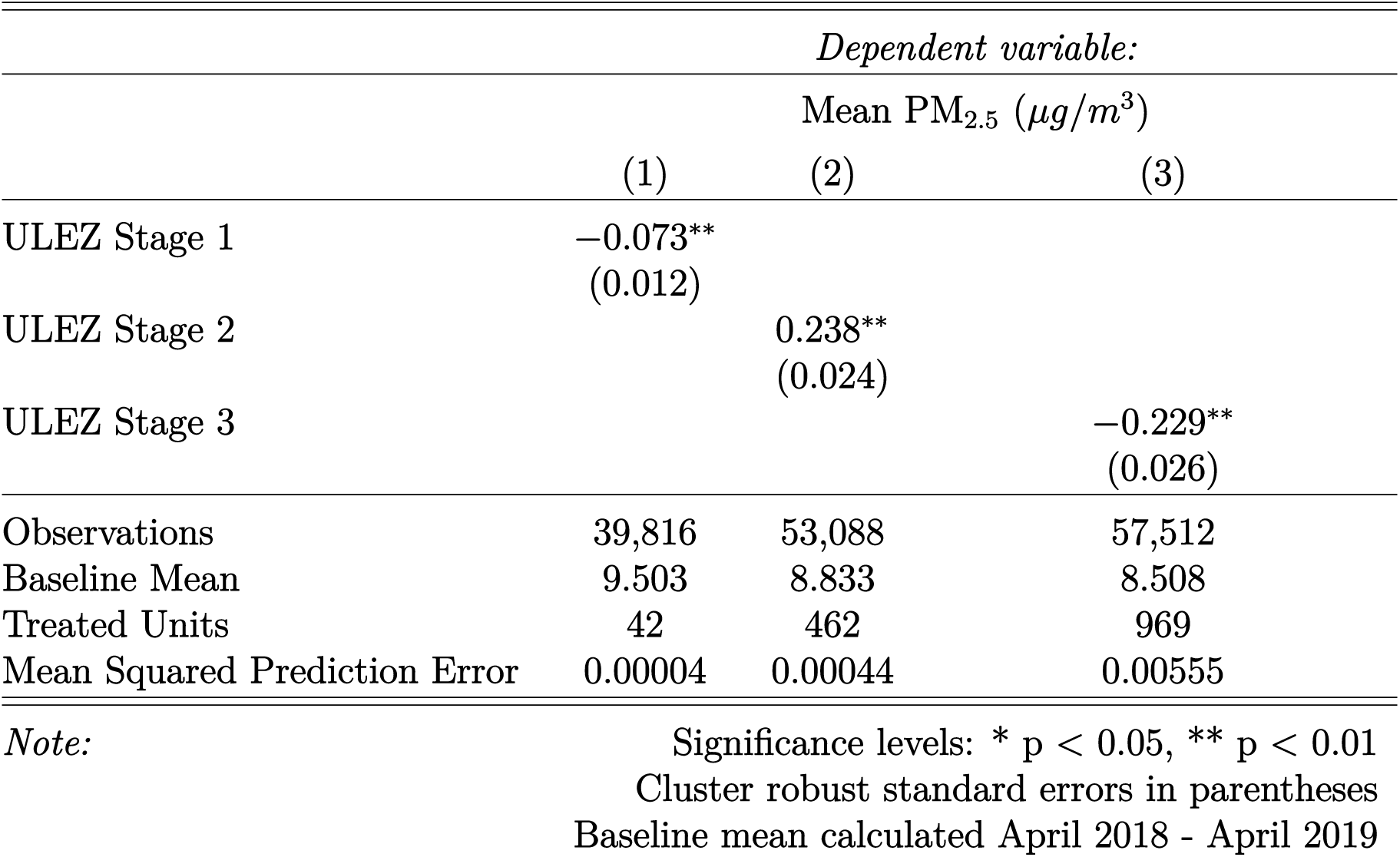
Synthetic Control Results - PM_2.5_ Levels.

|  | *Dependent variable:* |  |  |
| --- | --- | --- | --- |
| | Mean PM<sub>2.5</sub> ($\mu\text{g}/\text{m}^3$) | | |
|  | (1) | (2) | (3) |
| ULEZ Stage 1 | -0.073**<br>(0.012) |  |  |
| ULEZ Stage 2 |  | 0.238**<br>(0.024) |  |
| ULEZ Stage 3 |  |  | -0.229**<br>(0.026) |
| Observations | 39,816 | 53,088 | 57,512 |
| Baseline Mean | 9.503 | 8.833 | 8.508 |
| Treated Units | 42 | 462 | 969 |
| Mean Squared Prediction Error | 0.00004 | 0.00044 | 0.00555 |

*Note:*

We use the estimated reduction in PM_2.5_ levels to calculate the percentage reduction as a fraction of the initial baseline average. The results are displayed in Table 3.

**Table 3:** Percentage Change in PM_2.5_ levels by ULEZ Stage.

| ULEZ Stage | %$\Delta$ PM<sub>2.5</sub> |
| --- | --- |
| Stage 1 | 0.77% |
| Stage 2 | 2.69% |
| Stage 3 | -2.69% |

We find that the first ULEZ stage was associated with a minimal - but statistically significant - reduction in PM_2.5_ levels of 0.77% in the first 6 months. Paradoxically, in the second stage an initial increase of 2.69% was observed. In the final stage, an equivalent decrease of 2.69% was found in the first 4 months - the latest that could be observed due to limited data.

The extent of any spillover effect was also considered, to see if an improvement in pollution levels extended beyond the ULEZ boundary. Table 4 shows the effect of the first two stages on PM_2.5_ levels in the wider boundaries.

**Table 4:** Synthetic Control Results - Effect on PM_2.5_ Levels in Wider Practices.

|  | *Dependent variable:* |  |  |
| --- | --- | --- | --- |
| | Mean PM<sub>2.5</sub> ($\mu g/m^3$) | | |
|  | (1) | (2) | (3) |
| ULEZ Stage 1 | -0.073**<br>(0.012) | -0.018**<br>(0.002) | -0.026**<br>(0.005) |
| ULEZ Stage 2 |  | 0.238**<br>(0.024) | -0.087**<br>(0.024) |
| ULEZ Stage 3 |  |  | -0.229**<br>(0.026) |
| Observations | 39,816 | 53,088 | 57,512 |
| Baseline Mean | 9.503 | 8.833 | 8.508 |
| Treated Units | 42 | 462 | 969 |

*Note:*

**Table 5:** Percentage Change in PM_2.5_ Levels by ULEZ Stage and Boundary.

| ULEZ Stage | %Δ PM<sub>2.5</sub> |  |  |
| --- | --- | --- | --- |
|  | (1) | (2) | (3) |
| Stage 1 | **−0.77%** | −0.20% | −0.31% |
| Stage 2 | — | **2.69%** | −1.02% |
| Stage 3 | — | — | **−2.69%** |

*Note:* Numbers in parentheses indicate ULEZ boundary Values in bold represent results displayed in Table 3

We again see statistically significant but minimal changes to PM_2.5_ levels, implying there is very little spillover effect from improved pollution as a result of the ULEZ. This is in keeping with current literature, which found pollutants tend to disperse around 100-400 metres from their emission source [14].

### 3.2 Prescribing Outcomes

Applying the same method GCSM method, we construct a ‘synthetic London counter-factual’ from our weighted control units to estimate the effect of each ULEZ stage on bronchodilator and inhaled corticosteroid prescribing in the first 6 months after policy introduction. The results are shown in Tables 6 and 7.

**Table 6:**
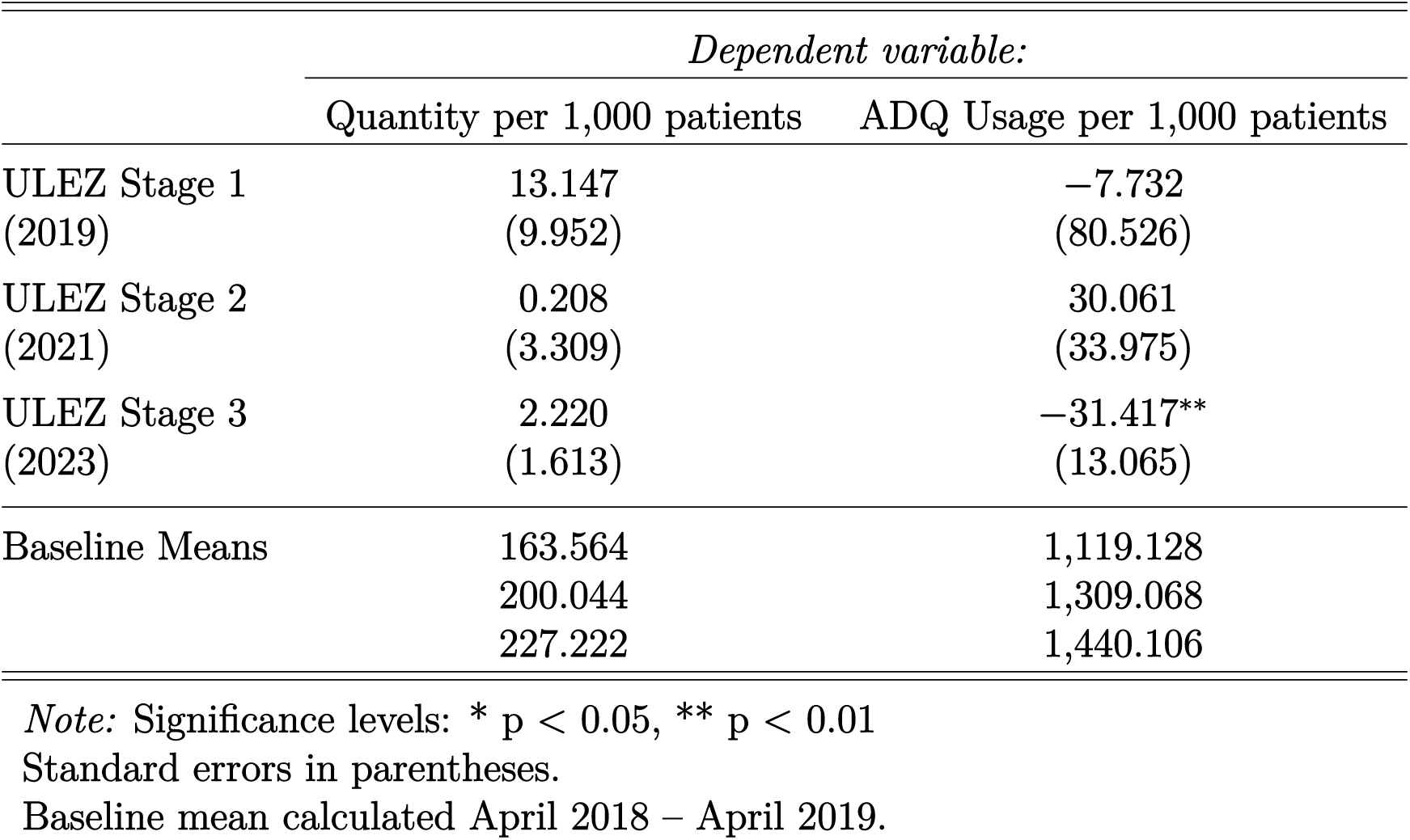
Estimated %Δ in bronchodilator prescriptions in Greater London in the 6 months following each ULEZ stage introduction.

|  | *Dependent variable:* |  |
| --- | --- | --- |
|  | Quantity per 1,000 patients | ADQ Usage per 1,000 patients |
| ULEZ Stage 1<br>(2019) | 13.147<br>(9.952) | −7.732<br>(80.526) |
| ULEZ Stage 2<br>(2021) | 0.208<br>(3.309) | 30.061<br>(33.975) |
| ULEZ Stage 3<br>(2023) | 2.220<br>(1.613) | −31.417**<br>(13.065) |
| Baseline Means | 163.564<br>200.044<br>227.222 | 1,119.128<br>1,309.068<br>1,440.106 |

*Note:* Significance levels: \* $p < 0.05$ , \*\* $p < 0.01$
Standard errors in parentheses.
Baseline mean calculated April 2018 – April 2019.

**Table 7:** Estimated %Δ in inhaled corticosteroid prescriptions in Greater London in the 6 months following each ULEZ stage introduction.

|  | *Dependent variable:* |  |
| --- | --- | --- |
|  | Quantity per 1,000 patients | ADQ Usage per 1,000 patients |
| ULEZ Stage 1<br>(2019) | −0.800<br>(0.415) | −19.785<br>(0.642) |
| ULEZ Stage 2<br>(2021) | −0.893**<br>(0.234) | 144.101**<br>(25.155) |
| ULEZ Stage 3<br>(2023) | 0.121<br>(0.363) | −11.678<br>(10.344) |
| Baseline Means | 15.526<br>20.113<br>22.282 | 803.978<br>1,151.958<br>1,258.414 |

*Note:* Significance levels: \* $p < 0.05$ , \*\* $p < 0.01$
Standard errors in parentheses.
Baseline mean calculated April 2018 – April 2019.

**Table 8:**
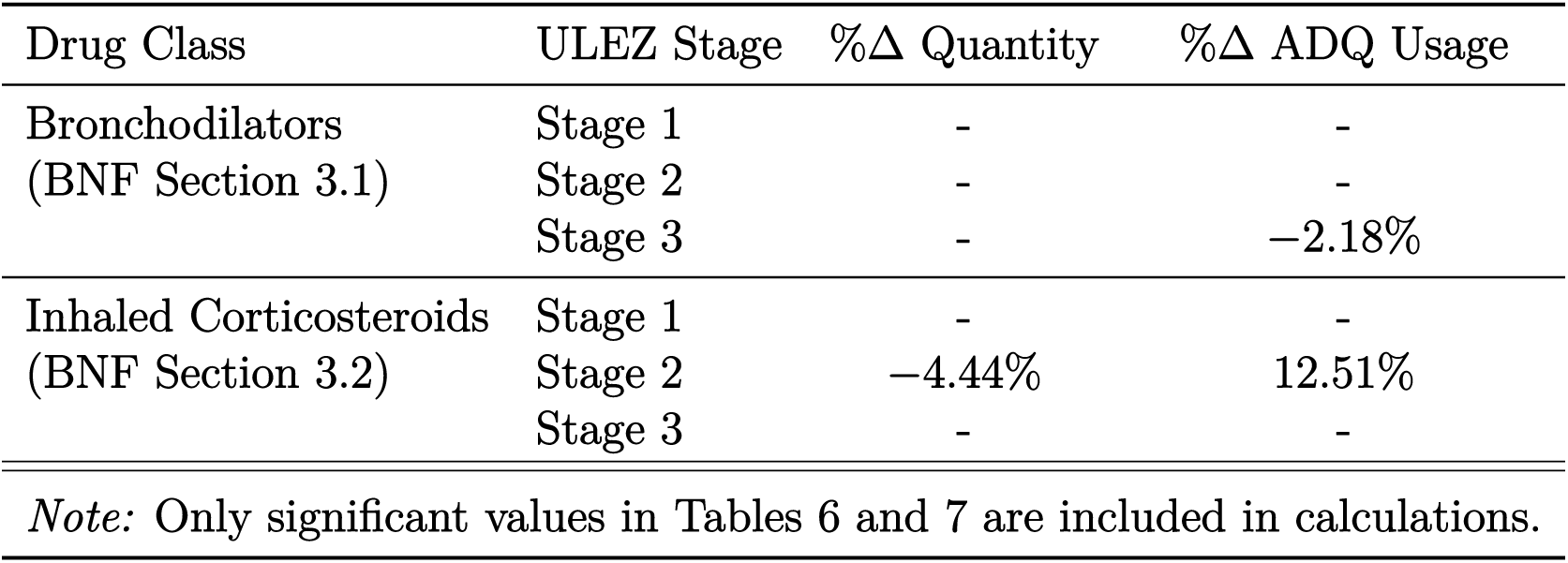
Percentage Change in Quantity and ADQ Usage per 1,000 patients by ULEZ Stage.

We find the introduction and subsequent expansions of the ULEZ have had mixed effects on respiratory prescribing trends. The first stage was found to have no significant effect on any metrics for either bronchodilator or inhaled corticosteroid prescribing. The second stage led to a 4.44% decrease in quantity of inhaled corticosteroids prescribed, and a 12.51% increase in ADQ usage. The final stage found there to be a 2.18% decrease in inhaled corticosteroid ADQ usage, but no other significant results. This is partially consistent with the PM_2.5_ trends observed.

### 3.3 Robustness Checks

#### 3.3.1 Placebo Tests

To ensure the robustness of the pollution results, placebo tests were performed by creating an artificial treatment identifier 6 months before each policy introduction. Table A4 shows these results.

The placebo tests found no statistically significant results for PM_2.5_ estimates, confirming the robustness of the results found and ensuring they are not attributed to wider downward pollution trends.

#### 3.3.2 BNF Section 20: Dressings

To ensure robustness of prescribing estimates, the analysis was repeated using prescribed items from BNF section 20: Dressings. As there is no plausible mechanism linking the ULEZ to prescribing patterns for dressings, we expected no significant change in quantity following the policy implementation. The results confirmed this expectation, supporting the validity of our main findings. This robustness check is presented in Table A5.

#### 3.3.3 Removing Cities

Multiple other English cities have introduced similar LEZs or clean air zones to reduce urban pollution levels; of the 18 control cities in this study, 6 have one such zone^3^. To ensure the robustness of our results, these cities were removed from the dataset and the analysis was performed again. They showed a similar pattern to our original results; modest decreases in PM_2.5_ levels in the first and third stages, and a small increase in the second stage. The results are shown in Table A6.

## 4 Discussion

### 4.1 PM_2.5_ Levels

This study found that the first ULEZ stage reduced PM_2.5_ levels by just 0.77% in the first 6 months, a negligible result in line with the findings of Ma, Graham and Stettler [15], and contrary to the findings of Beshir and Fichera [7]. In the second stage, we observed an increase in PM_2.5_ levels of 2.69% compared to the synthetic control, suggesting a paradoxical post-introduction effect similar to that observed by Park and Lim [16] in Seoul’s ULEZ. We surmise that this is likely partly due to the so-called ‘NO_X_ disbenefit effect’, whereby a short term decrease in NO_X_ can paradoxically cause PM_2.5_ levels to rise in the short term, suggesting the ULEZ improved air quality by other measures in the short run [17, 18]. Indeed it is a limitation of this study that satellite derived NO_X_ data is not available at an appropriate resolution to conduct an investigation, and so we cannot say with certainty if this is the cause of the increase in PM_2.5_. In the final stage, a decrease in PM_2.5_ levels of 2.69% was observed. This is equal in the negative direction to the second stage result at six months, and suggests that the final stage of the ULEZ was the most effective in reducing PM_2.5_ levels in the short term. However, the effect is still much smaller than the consensus value in the literature (*≈* 30% decrease). We hypothesise that overestimations of the effect of the ULEZ are caused by less sophisticated methods of analysis to establish a causal effect, as well as the use of more volatile and less comprehensive roadside monitoring data. Unfortunately, we cannot observe levels after 2023 due to data limitations, so the effects of the final stage past the initial four months cannot be assessed.

### 4.2 Prescribing Trends

We observed limited impacts of the ULEZ on primary care respiratory prescribing, broadly in line with our initial results estimating PM_2.5_ levels. The absence of significant prescribing changes following the first ULEZ stage follows the minimal change to air quality. Following the rise in PM_2.5_ levels after the second ULEZ stage, inhaled corticosteroid prescriptions declined in quantity yet increased substantially in ADQ usage. This divergence suggests that while fewer units were dispensed overall, patients already using inhaled corticosteroids required higher daily doses or moved toward more potent formulations; this is consistent with a worsening of underlying disease severity among the most vulnerable. Including the ADQ usage metric in this study allows us to elicit these trends more clearly than analysing prescription quantity alone. In the final stage, a small decrease in bronchodilator ADQ usage was observed, but no other respiratory prescribing metrics were affected. The final stage of the ULEZ showed a more substantial decrease in PM_2.5_ levels, and this provides limited evidence of a ‘knock-on’ effect to respiratory prescribing.

## 5 Conclusion

This study applied a generalised synthetic control model to estimate the impact of the three stages of London’s Ultra Low Emission Zone (ULEZ) on PM_2.5_ levels and cardiorespiratory prescribing trends. Contrary to existing literature on London’s ULEZ, we observed only minimal reductions in PM_2.5_ and prescribing activity.

While this study offers novel insight using advanced methods and new data, further research with more recent and individual-level data is needed. Overall, the ULEZ appears less effective at improving air quality and health outcomes than previously suggested, and likely requires integration with broader policy measures to have significant impact.

## Appendix: Tables

**Table A1:** Selected Control Areas and Sub-ICB Codes.

| Sub-ICB Code | Area |
| --- | --- |
| 09D | NHS Brighton and Hove |
| 36J | NHS Bradford District and Craven* |
| 15E | NHS Birmingham and Solihull* |
| B2M3M | NHS Coventry and Warwickshire |
| 01E | NHS Greater Preston |
| 15F | NHS Leeds |
| 04C | NHS Leicester City |
| 99A | NHS Liverpool |
| 14L | NHS Manchester |
| 13T | NHS Newcastle Gateshead* |
| 52R | NHS Nottingham and Nottinghamshire |
| 10R | NHS Portsmouth* |
| 01G | NHS Salford |
| 03N | NHS Sheffield* |
| 00P | NHS Sunderland |
| 02H | NHS Wigan Borough |
| 10Q | NHS Oxfordshire* |
| 26A | NHS Norfolk and Waveney |

*Note:* \* indicates a city within the ICB has a LEZ/clean air zone in place.

**Table A2:** Bronchodilators (BNF 3.1) with ADQ, Unit, and Administration Route.

| BNF Name | ADQ | Unit | Admin. Route |
| --- | --- | --- | --- |
| Aminophylline | 450 | mg | Oral |
| Bambuterol | 20 | mg | Oral |
| Eformoterol (Formoterol) | 24 | mcg | Inhal. powder |
| Ephedrine | 50 | mg | Oral |
| Fenoterol & ipratropium 100mcg/40mcg | 3 | puffs | Inhal. aerosol |
| Fenoterol & ipratropium 1.25mg/500mcg/4ml | 2 | vials | Inhal. solution |
| Indacaterol | 150 | mcg | Inhal. powder |
| Ipratropium | 0.12 | mg | Inhal. aerosol/powder |
| Ipratropium | 1.25 | mg | Inhal. solution |
| Isoprenaline | 0.64 | mg | Inhal. aerosol |
| Orciprenaline | 6 | mg | Inhal. aerosol |
| Orciprenaline | 60 | mg | Oral |
| Oxitropium | 0.4 | mg | Inhal. aerosol |
| Pirbuterol | 1.2 | mg | Inhal. aerosol |
| Pirbuterol | 30 | mg | Oral |
| Reproterol | 2 | mg | Inhal. aerosol |
| Rimiterol | 1.6 | mg | Inhal. aerosol |
| Salbutamol | 12 | mg | Oral |
| Salbutamol | 0.4 | mg | Inhal. aerosol/powder |
| Salbutamol | 10 | mg | Inhal. solution |
| Salbutamol CFC-free | 0.4 | mg | Inhal. aerosol |
| Salbutamol & ipratropium 20mcg/100mcg | 4 | puffs | Inhal. aerosol |
| Salbutamol & ipratropium 2.5mg/500mcg | 2 | vials | Inhal. solution |
| Salmeterol | 0.1 | mg | Inhal. aerosol/powder |
| Terbutaline | 20 | mg | Inhal. solution |
| Terbutaline | 1 | mg | Inhal. aerosol/powder |
| Terbutaline | 15 | mg | Oral |
| Theophylline | 400 | mg | Oral |
| Tiotropium | 5 | mcg | Inhal. Soln. |
| Tiotropium | 18 | mcg | Inhal. powder |

Source: [19].

**Table A3:** Corticosteroids (BNF 3.2) with ADQ, Unit, and Administration Route.

| BNF Name | ADQ | Unit | Admin. Route |
| --- | --- | --- | --- |
| Ciclesonide | 0.16 | mg | Inhal. aerosol |
| Beclometasone & salbutamol 50mcg/100mcg | 2 | puffs | Inhal. aerosol |
| Beclometasone & salbutamol 100mcg/200mcg | 4 | rotacaps | Inhal. powder |
| Beclometasone & salbutamol 200mcg/400mcg | 4 | rotacaps | Inhal. powder |
| Beclometasone | 0.4 | mg | Inhal. aerosol/powder |
| Beclometasone CFC-free (QVAR) | 0.2 | mg | Inhal. aerosol |
| Beclometasone and Formoterol 100mcg/6mcg | 4 | puffs | Inhal. aerosol |
| Budesonide | 1.5 | mg | Inhal. suspension |
| Budesonide | 0.4 | mg | Inhal. aerosol/powder |
| Budesonide & Formoterol | 2 | puffs | Inhal. aerosol/powder |
| Budesonide & Formoterol 100/6mcg, 200/6mcg | 2 | puffs | Inhal. aerosol/powder |
| Fluticasone | 0.2 | mg | Inhal. aerosol/powder |
| Fluticasone | 2 | mg | Inhal. suspension |
| Fluticasone & salmeterol (accuhaler) | 2 | puffs | Inhal. aerosol/powder |
| Fluticasone & salmeterol (evohaler) | 4 | puffs | Inhal. aerosol/powder |
| Mometasone | 0.4 | mg | Inhal. powder |

Source: [19].

### A.1 Synthetic Control Placebo

**Table A4:** Synthetic Control Placebo Results - PM_2.5_.

|  | *Dependent variable:* |  |  |
| --- | --- | --- | --- |
| | Mean PM<sub>2.5</sub> ($\mu g/m^3$) | | |
|  | (1) | (2) | (3) |
| ULEZ Stage 1 | -0.013<br>(0.019) | -0.004<br>(0.003) | -0.113<br>(0.010) |
| ULEZ Stage 2 |  | 1.184<br>(0.876) | -0.033<br>(0.017) |
| ULEZ Stage 3 |  |  | 0.006<br>(0.004) |
| Observations | 39,816 | 53,088 | 57,512 |
| Baseline Mean | 9.503 | 8.833 | 8.508 |
| Treated Units | 42 | 462 | 969 |
| Mean Squared Prediction Error | 0.00004 | 0.00044 | 0.00555 |

*Note:* Significance levels: \* $p < 0.05$ , \*\* $p < 0.01$ Cluster robust standard errors in parentheses Baseline mean calculated April 2018 - April 2019

**Table A5:** Synthetic Control Results – Dressings.

|  | *Dependent variable:* |  |
| --- | --- | --- |
|  | Quantity | ADQ Usage |
| ULEZ Stage 1 | 19.823<br>(10.552) | —<br>— |
| ULEZ Stage 2 | -2.950<br>(3.131) | —<br>— |
| ULEZ Stage 3 | 1.632<br>(2.811) | —<br>— |

*Note:* Significance levels: \* $p < 0.05$ , \*\* $p < 0.01$ Standard errors in parentheses.
*Note:* ADQ values are not supplied for dressings.

**Table A6:** Robustness Test Results - Removing Cities.

|  | *Dependent variable: Mean $PM_{2.5}$ ($\mu g/m^3$)* |  |  |
| --- | --- | --- | --- |
|  | (1) | (2) | (3) |
| ULEZ Stage 1 | −0.069**<br>(0.011) |  |  |
| ULEZ Stage 2 |  | 0.122**<br>(0.032) |  |
| ULEZ Stage 3 |  |  | −0.015*<br>(0.006) |

*Note:* Significance levels: \* $p < 0.05$ , \*\* $p < 0.01$ Cluster robust standard errors in parentheses Baseline mean calculated April 2018 - April 2019

### A.2 Generalised Synthetic Control Method (GSCM)

#### A.2.1 Introduction

The Generalised Synthetic Control Method (GSCM), introduced by Xu [13], extends the standard SCM to address limitations in causal inference. Unlike the traditional SCM, which limits analysis to a single treated unit and assumes a fixed weighted average for counterfactual estimation, GSCM uses an interactive fixed effects model; allowing for multiple treated units and accounting for unobserved time-varying confounders.

#### A.2.2 Model Specification

Consider panel data with units *i* = 1, …, N over time periods *t* = 1*, …, T*. The observed outcome is given by:

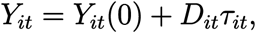

where:

- *Y_it_*(0) is the counterfactual outcome.
- *D_it_* is an indicator function, where *D_it_* = 1 if unit *i* is treated at time *t*, and *D_it_* = 0 otherwise.
- *ô_it_* is the treatment effect of interest.

#### A.2.3 Interactive Fixed Effects Model

GSCM models the counterfactual outcome as:

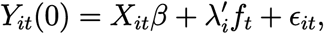

where:

- *X_it_* represents observed covariates.
- *â* is the vector of associated coefficients.
- *ë^S^ f_t_* captures unobserved time-varying confounders via interactive fixed effects.
- *ɛ_it_* is an idiosyncratic error term.

The term *λ^S^ f_t_* allows for heterogeneous, time-varying unobserved factors, which the traditional SCM cannot accommodate.

#### A.2.4 Estimation of Counterfactuals

Since *λ_i_* and *f_t_* are unobserved, they are estimated from pre-treatment data using matrix completion [20]. The estimated counterfactual outcomes for treated units post-treatment are given by:

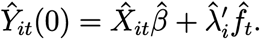

The treatment effect is then computed as:

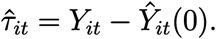

## Appendix: B Data Bibliography

### Prescribing Data

NHS Digital. *English Prescribing Dataset*. 2024. url: https://digital.nhs.uk/data-and-information/publications/statistical/english-prescribing-data (visited on 10/12/2024)

### PM_2.5_ Estimates

Siyuan Shen et al. ‘Enhancing Global Estimation of Fine Particulate Matter Concentrations by Including Geophysical *a Priori* Information in Deep Learning’. In: *ACS ES&T Air* 1.5 (10th May 2024), pp. 332–345. issn: 2837-1402, 2837-1402. doi: 10.1021/acsestair. 3c00054. url: https://pubs.acs.org/doi/10.1021/acsestair.3c00054 (visited on 10/12/2024)

### Demographic Control Variables

Public Health England. *PHE Fingertips Health Profiles*. 2024. url: https://fingertips.phe.org.uk (visited on 07/11/2024)

### Quality and Outcomes Framework (QOF)

NHS England Digital. url: https://digital.nhs.uk/data-and-information/data-tools-and-services/data-services/general-practice-data-hub/quality-outcomes-framework-qof (visited on 21/06/2025)

### GP list and Geographic Information

NHS Digital. *English Prescribing Dataset*. 2024. url: https://digital.nhs.uk/data-and-information/publications/statistical/english-prescribing-data (visited on 10/12/2024)

NHS Digital. *GP and GP Practice Related Data*. 2024. url: https://digital.nhs.uk/services/organisation-data-service/export-data-files/csv-downloads/gp-and-gp-practice-related-data (visited on 01/10/2024)

### ULEZ Boundary Postcodes

Transport for London. *\*FOI Request on ULEZ Expansion Consultation Data* [Freedom of Information Request].* FOI-1327-2223. 2023. url: https://tfl.gov.uk/corporate/transparency/freedom-of-information/foi-request-detail?referenceId=FOI-1327-2223

Transport for London. *\*FOI Request on ULEZ Postcode List* [Freedom of Information Request].* FOI-2281-2223. 2023. url: https://tfl.gov.uk/corporate/transparency/freedom-of-information/foi-request-detail?referenceId=FOI-2281-2223

## Data Availability

All data analysed in the present study is available online at the sources listed in the data bibliography or data availability links.

https://digital.nhs.uk/dataand-information/publications/statistical/english-prescribing-data

https://fingertips.phe.org.uk

https://digital.nhs.uk/data-and-information/data-tools-and-services/data-services/general-practice-data-hub/quality-outcomes-framework-qof

https://digital.nhs.uk/data-and-information/publications/statistical/english-prescribing-data

https://digital.nhs.uk/data-and-information/publications/statistical/english-prescribing-data

https://tfl.gov.uk/corporate/transparency/freedom-of-information/foi-request-detail?referenceId=FOI-1327-2223

https://tfl.gov.uk/corporate/transparency/freedom-of-information/foi-request-detail?referenceId=FOI-2281-2223

## Footnotes

1 Due to data limitations, only the first 4 months post-policy introduction is estimated for Stage 3.

2 These were filtered according to sub-ICB location, ensuring that surrounding suburban areas were included too.

3 These are: Bradford, Birmingham, Newcastle, Oxford, Portsmouth and Sheffield.

